# Immunotherapy Implication of Signature-Guided Biomarker Discovery for Trastuzumab-Resistant HER2-Positive Breast Cancer

**DOI:** 10.1101/2020.01.17.20017947

**Authors:** Andrea Sand, Aspen T. Duffin, Geoffrey T. Riddell, Mitchel Piacsek, Brittany Last, Chaoyang Sun, Richard A. Rovin, Judy A. Tjoe, Jun Yin

## Abstract

Despite the great improvement of patient outcomes by trastuzumab, a monoclonal antibody targeted on HER2-positive breast cancer, approximately 23% of patients with early-stage disease treated with adjuvant trastuzumab either fail to respond or experience recurrence within 10 years, highlighting the importance of identifying which HER2-positive patients would benefit from trastuzumab upfront. Efforts to identify biomarkers predictive of response to trastuzumab in initial breast tumor core biopsies have been complicated by the clinical and biological heterogeneity of HER2-positive tumors. Therefore, we identified a trastuzumab-resistant (TrR) signature that accurately predicts response to trastuzumab quantitively and qualitatively in vitro and in vivo, via repurposing transcriptome profiles in an engineered cell line model. We additionally demonstrated that our TrR signature was associated with tumor progression and capable of stratifying patient prognosis. Our study further illustrated the possible mechanism of this resistance as being less inherited cytotoxic T cell infiltration and failure to secrete Interferon-γ upon trastuzumab treatment in TrR tumors. These findings highlight the potential clinical application of TrR signature in treatment management and identifying possible immunotherapy interventions.

## Introduction

Breast carcinomas with HER2/ERBB2 receptor amplification and overexpression account for approximately 20% of all breast cancers, which also confer more aggressive phenotypes and are associated with poor prognosis (Arteaga, Sliwkowski et al., 2011, Singh, Jhaveri et al., 2014, Slamon, Clark et al., 1987). The clinical management of this cohort often relies on diagnosis and targeting of a single molecule, such as mutation status, amplification or gain/loss of function of HER2 (6). The strategy to target the HER2 receptor with a monoclonal blocking antibody (mAb), trastuzumab, has led to great improvement in overall survival in HER2-positive breast cancer. However, approximately 25% patients in this cohort still fail to respond to this treatment initially and almost 23% of patients with early-stage HER2-positive breast cancer treated with adjuvant chemotherapy and trastuzumab experience disease recurrence within 10 years (Perez, Romond et al., 2011, Perez, Romond et al., 2014), highlighting the importance of identifying which HER2-positive patients will benefit from this treatment and those that will not. Although other HER2-targeted agents, including lapatinib, pertuzumab, and trastuzumab emtansine (T-DM1), administered either in place of or in combination with trastuzumab have demonstrated activity in trastuzumab-resistant (TrR) cancers, the challenge remains to identify patients who require the addition of these agents upfront. Efforts to identify biomarkers in the initial breast tumor core biopsy predictive of response to trastuzumab have been complicated by the clinical and biological heterogeneity of HER2-positive tumors. The predictors generated from patient data are usually accompanied by insufficient sample size, heterogeneity in mutation status and treatment complexity; therefore, the conclusion usually remains elusive (Harris, You et al., 2007, Varadan, Gilmore et al., 2016). Exploring the transcriptome profiles of engineered cell line models to identify biomarkers for trastuzumab resistance could be one of the most promising approaches to overcoming existing challenges with their homogenous genetic background and drug specificity.

Increasing evidence that the immune system plays a significant role in the therapeutic effects of HER2-targeted therapy has been provided by multiple studies (Galluzzi, Buque et al., 2017, Gennari, Menard et al., 2004, Loibl, de la Pena et al., 2017, Muller, Kreuzaler et al., 2015, Musolino, Naldi et al., 2008, Stagg, Loi et al., 2011, Varadan et al., 2016, Varchetta, Gibelli et al., 2007), e.g., trastuzumab was able to facilitate killing tumor cells via antibody-dependent cellular cytotoxicity (ADCC) through recruitment of natural killer cells (NK cells)(Herrmann, Lehr et al., 2004, Okita, Mougiakakos et al., 2012). In addition, the presence of tumor-infiltrating lymphocytes (TIL) was also found to be associated with improved outcomes with trastuzumab treatment in clinical trials (Adams, Gray et al., 2014, Ibrahim, Al-Foheidi et al., 2014, Loi, Michiels et al., 2014, Loi, Sirtaine et al., 2013, Luen, Salgado et al., 2017, Perez, Ballman et al., 2016, Salgado, Denkert et al., 2015, Stanton & Disis, 2016). Lately, multiple clinical trials targeting the TrR cohort by modulation of certain cytokines, e.g., interleukin-2, interleukin-6, have been initiated based on preclinical studies (Mani, Roda et al., 2009, Repka, Chiorean et al., 2003, Zhong, Davis et al., 2016), suggesting that the interaction of tumor-secreted cytokines with innate or adaptive cells contributes to trastuzumab efficacy. However, other than NK cells, no study has yet correlated other types of immune cells with patient response to trastuzumab treatment.

In this study, we developed a 43-gene signature that can accurately predict the response to trastuzumab *in vitro* and *in vivo* by repurposing a publicly available transcriptome profile (GSE15043) (Gu, Waliany et al., 2009) via a different algorithm. We have further demonstrated its efficacy on stratifying patients who will benefit from trastuzumab treatment quantitively and qualitatively. The network of genes in our TrR signature indicates a novel mechanism that inherited less cytotoxic T cell infiltration in TrR tumors and failure of interferon gamma (IFN-γ) secretion upon trastuzumab treatment are likely responsible for treatment resistance. We further explored clinical relevance of this mechanism using our in-house treatment naïve patient samples. In summary, we identified a gene signature, which predicts clinical outcomes and further demonstrated a novel mechanism and its associated targetable key players.

## Results

### Current clinical biomarkers for trastuzumab sensitivity in HER2-positive breast cancer lack sufficient accuracy

Current treatment management of HER2-positive breast cancer often only relies on features of a single gene, such as mutation status, amplification or gain/loss of function of HER2 (Arnaout, Dawson et al., 1992, Centis, Tagliabue et al., 1992, Dowsett, Cooke et al., 2000). However, the growing population of primary or acquired resistance to trastuzumab in breast cancer patients with HER2 aberration indicates that current clinical makers such as HER2 mRNA, protein or activity levels for trastuzumab treatment still await improvement. To test this hypothesis in our study, two pairs of TrS (ZR75-30, SKBR3) and TrR cell lines (HCC1954, T47D) were accessed for their HER2 protein expression levels and their responses to trastuzumab treatment. Interestingly, HCC1954, which possesses a high expression levels of HER2, failed to respond to trastuzumab treatment at a fairly high dosage (Fig. 1A, top panel). In addition, we further analyzed the accuracy of HER2 protein levels, mRNA levels and phospho-HER2 (p-HER2) levels when predicting trastuzumab sensitivity across 15 HER2-positive breast cancer cell lines utilizing data from Cancer Cell Line Encyclopedia (CCLE) (Barretina, Caponigro et al., 2012) and Reverse Phase Protein Array (RPPA). Although p-HER2 has been reported to be a more promising marker than HER2 protein levels (accuracy rate of 65%) and HER2 mRNA levels (accuracy rate of 55%), its prediction accuracy only reached 69% (Fig. 1A, bottom panel).

**Figure 1.**
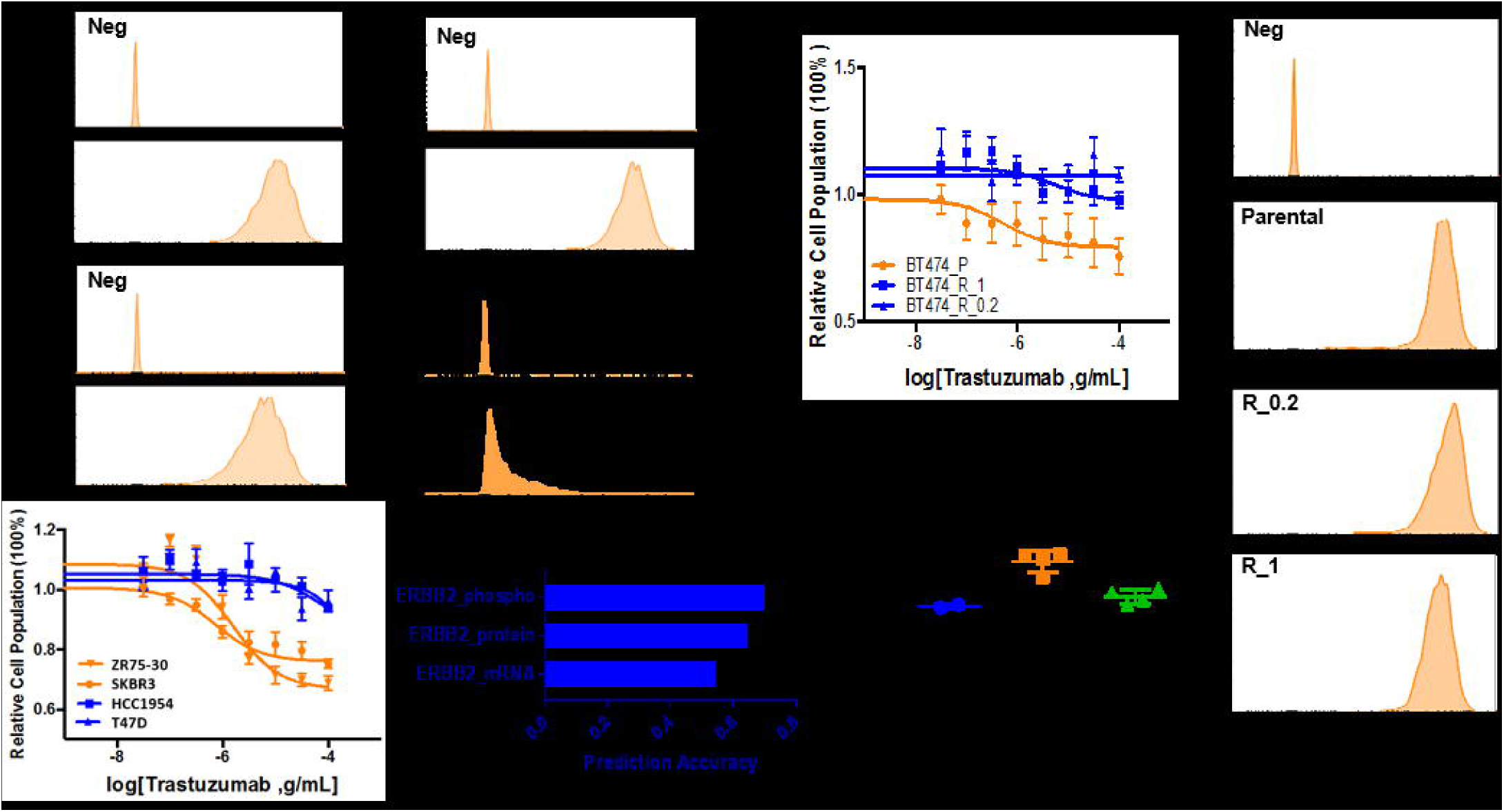
Current biomarkers for clinical management in treating HER2 (ERBB2)-positive breast cancer lack prediction accuracy. (A) The accuracy of biomarkers often used clinically, such as mRNA levels (labeled as ERBB2_mRNA), protein levels (labeled as ERBB2_protein) or activity levels of HER2 (indicated by ERBB2_phospho) to predict trastuzumab sensitivity amongst HER2-positive breast cancer cells are 0.55, 0.65 0.69, respectively, where 1 represents the highest accuracy. Two pairs of TrS (ZR75-30, SKBR3) and TrR cell lines (HCC1954, T47D) were tested for their trastuzumab sensitivity. (B) The isogenic cell line panel consisting of BT474 and BT474-derived TrR cell lines in trastuzumab at the concentration of 0.2mM (BT474_R_0.2) and 1Mm (BT474_R_1) were tested for HER2 protein expression levels via flow cytometry (right panel), mRNA levels via qPCR (bottom, left panel) and trastuzumab sensitivity (top, left panel).

In order to develop a set of biomarkers which are specific to trastuzumab sensitivity, we first established an isogenic panel of cell lines derived from BT474, a trastuzumab sensitive cell line. This panel included BT474 and two resistant clones of BT474 continuously cultured in 1uM or 0.2uM trastuzumab for three weeks (BT474_R_1 and BT474_R_0.2, Fig. 1B, top right panel). To evaluate whether this adapted resistance is associated with downregulation of HER2 levels, we further assessed their HER2 protein and mRNA levels but failed to detect any significant difference (Fig. 1B). Our observation indicated that the possible mechanism of TrR in this model was not dependent on HER2, indicating that HER2 alone as a biomarker is insufficient for clinical treatment decision.

### TrR signature accurately predicts trastuzumab sensitivity in cell lines and patient tumors

As gene expression data has proven to have a robust capacity for predicting drug sensitivity (Costello, Heiser et al., 2014), we identified a TrR signature consisting of 43 differentially regulated genes between BT474 and its derived TrR cell lines by leveraging their transcriptomic data, with the supervised clustering across the isogenic cell lines shown in Fig. 2A. To test the predictive power of TrR signature in vitro, we assessed the trastuzumab sensitivity data of 15 HER2-positive breast cancer cell lines via Cancer Therapeutics Response Portal (CTRPv2) (Basu, Bodycombe et al., 2013) and calculated their TrR scores from their microarray and RNA sequencing available at CCLE as a testing dataset. The efficacy of this signature yielded AUC values of 0.875 and 0.750 when calculated through RNA sequencing and microarray, respectively (Fig. 2B, top panel), suggesting our signature is rather sensitive to the profiling technique. To investigate if the TrR signature is relevant in patient cohorts, we applied our signature to publicly available dataset GSE62327 (Triulzi, De Cecco et al., 2015) and GSE55348 (Castagnoli, Iezzi et al., 2014) to test the ability of the TrR signature to identify patients within the HER2 breast cancer subtype who would less likely benefit from trastuzumab treatment. Our results suggest an accurate prediction of initial nonresponders to HER2 inhibition by trastuzumab within this cohort, indicated by an AUC of 0.843. Although the TrR signature was not designated to predict patients who would likely develop a relapse after an initial response to trastuzumab, our analysis demonstrated an accuracy of 82% when predicting this population, with an AUC of 0.714 (Fig. 2B). We further assessed TrR signature scores in both cell lines and patient tumors and demonstrated significant enrichments in positive signature scores in resistant cell lineages or tumors (Fig. 3C). Overall, our results indicate that this TrR signature could be a powerful tool to predict initial response to trastuzumab in vitro and in vivo, qualitatively and quantitively.

**Figure 2.**
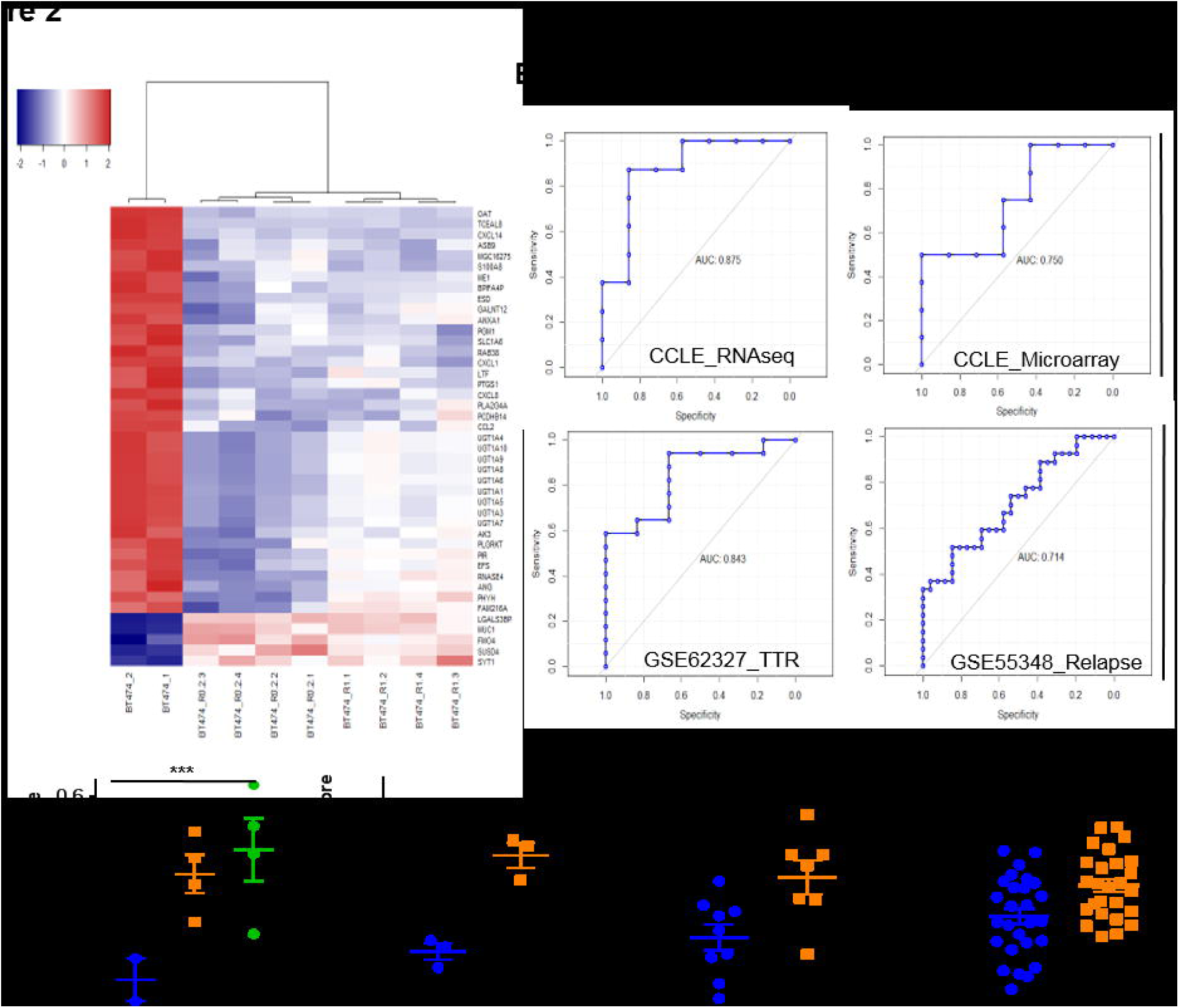
TrR signature predicts response to trastuzumab sensitivity in cell lines and patient tumors. (A) Supervised clustering by significantly differentially regulated 43 genes between BT474 and its derived TrR cell lines across the isogenic cell line panel. (B) Receiver-operating characteristic (ROC) curves for the prediction of training sets for trastuzumab sensitivity in breast cancer cell lines (top left, CCLE mRNA data; top right RNAseq data) and breast cancer patient tumors (bottom left, GSE62327; bottom right, GSE55348). Area under the curve (AUC) values were used as an indicator for accuracy. A ROC AUC value of 1 represents perfect prediction and 0.5 represents random chance. (C) TrR signature scores of BT474, its corresponding TrR lines, BT474 with control siRNA and HER2 siRNA, a training set of HER2-positive breast cancer cell lines with available data in CCLE and patient tumors with available data in GSE55348 were assessed and cross-compared.

**Figure 3.**
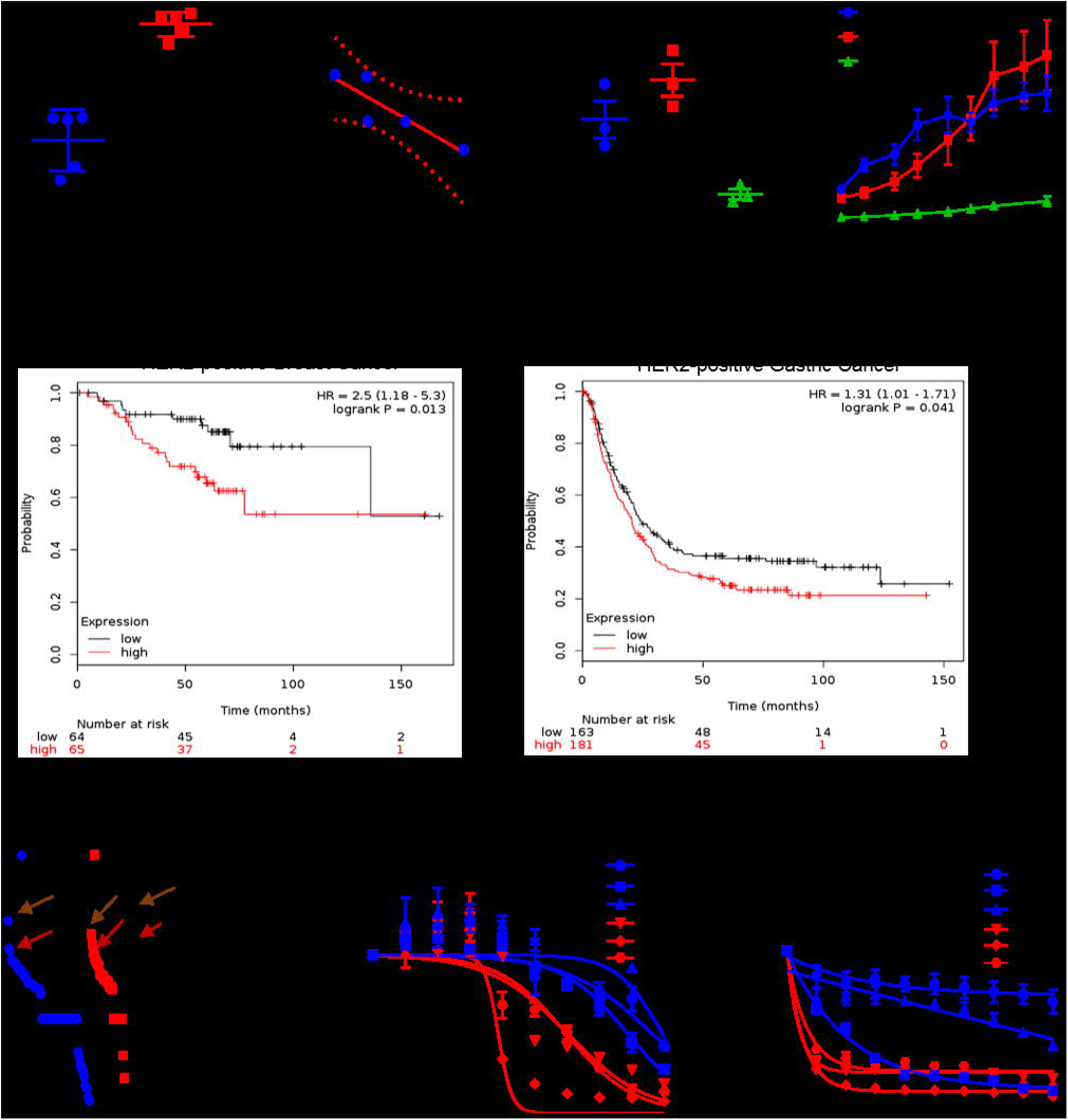
TrR signature predicts tumor progression, patient prognosis and is indicative of treatment for TrR patients. (A) TrR signature scores of five TrS cell lines and TrR cell lines were assessed and cross-compared (**P < 0.01). Signature scores and doubling time of cell lines which possess available data were correlated. (B) TrR signature scores of tumors dissected from patient-derived xenografts (PDX) generated from three TrR cell lines (BT474_R, HCC1954 and T47D) were assessed and cross-compared (**P < 0.01, left panel) and the corresponding tumor growth curves were exhibited and cross-compared (right panel). (C) Patient overall survival analysis in HER2-positive breast cancer and HER2-positive gastric cancer that were treated with trastuzumab were separated based on TrR signature scores. log-rank p-value is displayed that shows statistical significance. (D) Drug candidates available in Connectivity Map database, which include gene expression profiles with or without approximately 100 drug perturbation in PC3, HL60 and MCF7 cell lines, were sorted based on their predicted scores. A score close to 1 represents the highest likelihood of its effectiveness on targeting TrR cells (left panel). Viability curves following treatment of the top two drug candidates, Trichostatin A (brown arrows) and Geldanamycin (red arrows), for five days in three TrS (BT474, SKBR3 and ZR75-30) cells and three TrR cells (BT474_R, HCC1954 and T47D).

### TrR signature predicts tumor progression, patient overall survival and TrR tumor-targeting agents

Within the 43 genes in the TrR signature, we have detected and validated amplification of certain oncogenes (e.g., MUC1, Fig. S1), which are proven to be associated with tumor growth and progression (Horm, Bitler et al., 2012, Pochampalli, Bitler et al., 2007). Interestingly, we found a significant negative correlation (correlation coefficient of −0.86) between TrR signature scores and doubling time within five different HER2-positive breast cancer cell lines (Fig. 3A). To test whether this observation applies in vivo, we appraised TrR signature scores through qPCR and growth curves of three TrR cell line (BT474_R, HCC1954, T47D) – derived xenografts. It was notable that T47D, which exhibited the slowest tumor progression rate, had the lowest TrR signature score (Fig. 3B). HCC1954, which had the steepest exponential growth rate and largest tumor size at the end point of the experiment, exhibited the highest TrR score. To validate this signature in patient cohorts, we tested the performance of TrR signature in a HER2-positive breast cancer cohort (n =220) and HER2-gastric cancer cohort (n = 453), both of which had trastuzumab as their first-line treatment (Digklia & Wagner, 2016). We report that our TrR signature was able to stratify patient survival in both cohorts, suggesting that our signature is insensitive to different cancer types with HER2 amplification but rather sensitive to the treatment itself. Taken together, these results demonstrate that the TrR signature accurately stratifies tumor progression in vitro and in vivo, as well as patient prognosis following trastuzumab treatment.

Given that TrR signature is associated with tumor progression, we asked whether we could identify agents that would reverse the TrR signature and thereby reduce the growth of TrR cells. To this end, we compared data from Connectivity Map (CMAP) with TrR signature. The CMAP is a public database with many drug-associated gene expression profiles (Lamb, Crawford et al., 2006). Remarkably, after searching for agents that could reverse gene expression profiles in the TrR signature and therefore might be expected to impede cell growth, we found that HDAC inhibitor, Trichostatin A, and Hsp90 inhibitor, Geldanamycin were ranked near the top of the CMAP list in terms of reversing TrR signature (brown arrows, Fig. 3D, left panel). Furthermore, we validated the efficacy of Trichostatin A and Geldanamycin and found that they specifically target TrR cells (Fig. 3D, middle and left panel), which is consistent with outcomes from other studies (Damaskos, Garmpis et al., 2017, De Mattos-Arruda & Cortes, 2012). The ability of the TrR signature to efficiently identify drugs that target TrR tumors provides evidence that the TrR signature is linked to biological functions involved in tumor progression.

### TrR signature reveals novel mechanism of TrR resistance

The innate resistance or adaptive resistance to trastuzumab is a combined effect of co-mutations/co-genetic alternations in cancer cells, which could be more determinative than the effects of individual alterations. Here, we hypothesize that the network of 43 genes in the TrR signature, with the top predicted canonical pathways being inhibited cytokine-cytokine receptor interaction, inhibited immune cell trafficking etc., indicates the mechanism of TrR resistance (Fig. 4A). We further investigated which immune cell type might be associated with this resistance by collecting seven different immune cell signatures and correlated TrR signature scores of our isogenic cell line panel derived from BT474 with scores calculated from the seven signatures. As shown in Fig. 4B, cytotoxic T-cell signature exhibited an extremely similar performance in distinguishing BT474 from its derived TrR cell lines with TrR signature, with a correlation coefficient as high as 0.61. Based on this observation, we performed immunohistochemistry staining of CD4 and CD8, markers that are commonly used to identify cytotoxic T cells (Zhang & Bevan, 2011), on our in-house FFPE samples (core biopsies, treatment naïve) of patients whose long term clinical outcomes proved to be sensitive or resistant to trastuzumab treatment. Significantly greater infiltration of CD8+/CD4+ T cells were observed in TrS patients, regardless of their ER status, shown in Fig. 4C. To determine whether this infiltration is associated with tumor-secreted cytokines or chemokines as predicted in the pathway analysis, we performed ELISA cytokine arrays on culture media secreted by BT474 and BT474_R with or without trastuzumab treatment. Interestingly, significantly higher levels of multiple cytokines associated with cytotoxic T-cell recruitment and infiltration (CCL5, ICAM-1) (Maimela, Liu et al., 2019) were detected in BT474-secreted media (Fig. 4D), which is consistent with the observation of more infiltrated T cells in TrS patient samples. We also observed a strong stimulation of IFN-γ and CD40L secretion, which are commonly associated with cytotoxic functions of CD8+ T cells through the production of TNF-related apoptosis-inducing ligands, ROS (McClory, Hughes et al., 2012) and Perforin (Wongtrakoongate, 2015), in BT474-containing media with trastuzumab treatment. In contrast, inhibited secretion of those two cytokines was detected in BT474_R-containing media post-trastuzumab treatment. Overall, our results suggest that deficiency of cytotoxic T cell infiltration due to insufficient secretion of CCL5 or ICAM-1 in tumors of TrR patients, combined with failure to stimulate tumor cells to secrete IFN-γ and CD40L by trastuzumab treatment, may be a possible mechanism contributing to trastuzumab resistance in TrR tumors.

**Figure 4.**
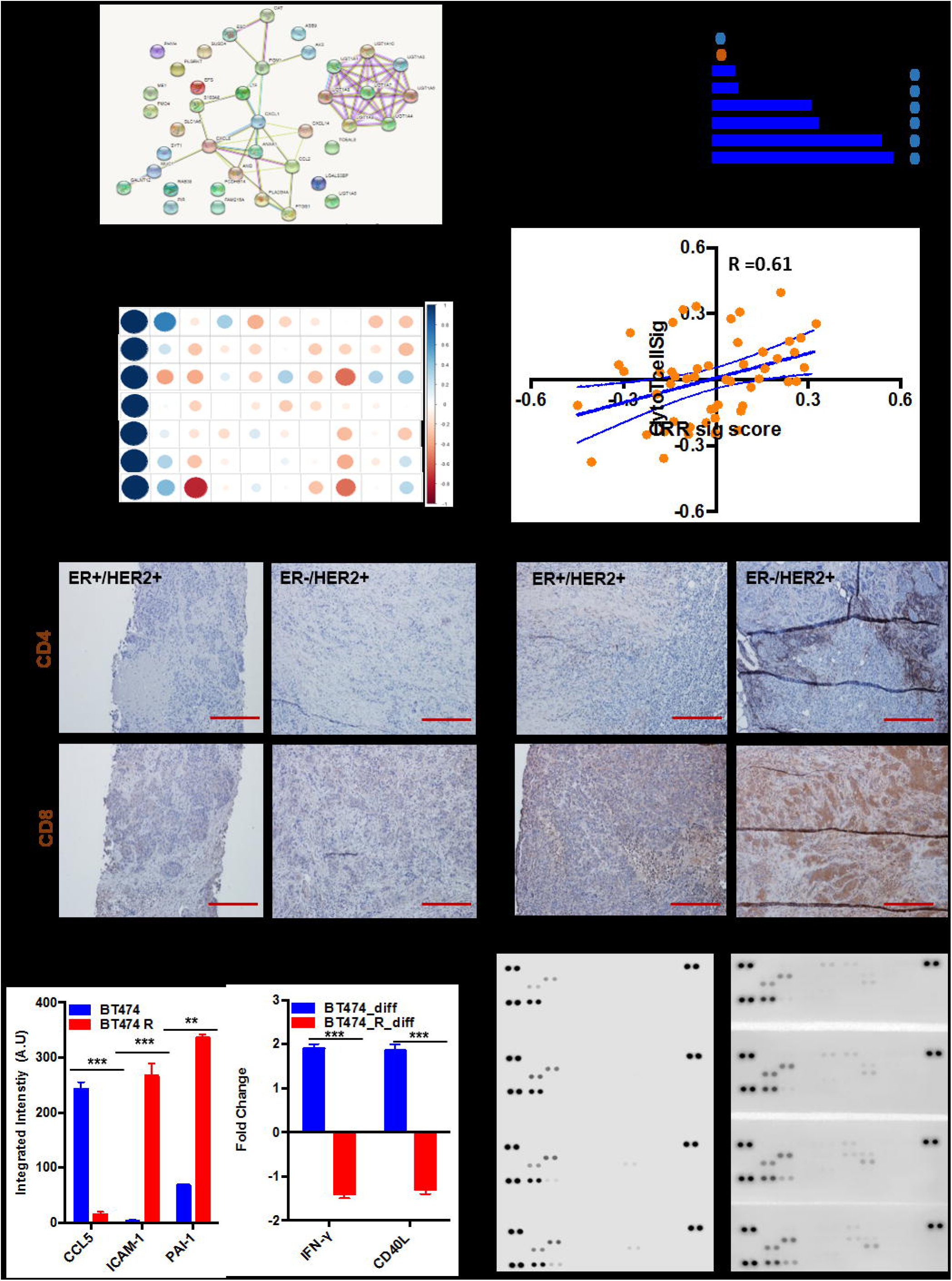
Network of gene regulators in TrR signature reveals possible mechanism of TrR resistance. (A) Network of gene regulators in TrR signature (left panel) and the top canonical pathways involving those regulators are exhibited (right panel). (B) Correlation coefficient from linear regression between TrR signature scores and a series of other available innate immune cells’ signatures (cytotoxic T-cell signature, B cell signature, T-cell signature, immunology index, monocyte signature and myeloid signature) across BT474 and its derived TrR cells lines. (C) Immunohistology staining of CD4 and CD8 on in-house TrR and TrS HER2-positive breast cancer patient FFPE samples (scale bar: 200μm). (D) Significantly differentially secreted cytokine amongst media secreted by BT474 and BT474_R with and without 1μM trastuzumab treatment (right panel, **P<0.01, ***P<0.001), detected by cytokine ELISA array with representative blots (left panel).

## Discussion

HER2 immunoreactivity of HER2-positivie breast cancers are often used clinically for treatment management. Targeting of the HER2 receptor in the form of a monoclonal blocking antibody, such as trastuzumab, has shown great improvement in patient survival. However, a large population of non-responders to this treatment and a recurrence rate of 23% for HER2-positive breast cancer patients treated with trastuzumab has highlighted the importance of identifying biomarkers which can accurately predict response or resistance to anti-HER2 therapy with more complex mechanism of action which cannot be represented by a single molecular marker.

Intriguingly, transcriptomic analysis, which relies on robust gene expression signatures designed to capture the core common features indicative of drug sensitivity, regardless of their precise molecular origin, represents one of the most promising approaches to overcome the current challenge. Despite substantial efforts towards identify reliable biomarkers that predict benefits of trastuzumab treatment using patient tumors, those biomarkers usually only work well within a subpopulation in this cohort, e.g., a subtype with a particular molecular features (Varadan et al., 2016) rather than an entire cohort. The majority of these patient studies suffer from insufficient sample size, have various mutation status and treatment complexity within patient tumors. Therefore, leveraging transcriptomic data from appropriately engineered cell lines, which possess the same genetic background, can be one of the best representative proxies to identify therapeutic interventions since this model provides sufficient sample size, a cleaner background and is more drug-specific. In addition, the rapidly decreasing price of gene expression microarrays and RNAseq makes it plausible to integrate this technology as a companion diagnosis into clinical trials for the eventual individualization of patient care. Here, we identified a 43-gene TrR signature via a panel of engineered cell lines and demonstrated its high sensitivity and specificity in predicting response to trastuzumab *in vitro* and *in vivo*. Interestingly, our TrR signature is also able to stratify tumor progression and patient prognosis, therefore serving as a powerful tool to predict the efficacy of targeted interventions within this cohort. The efficacy of the predicted top two candidates, a HDAC inhibitor, Trichostain A, and a Hsp90 inhibitor, Geldanamycin, on specifically targeting TrR cell lines were validated. This observation is also consistent with outcomes of other studies. Our results indicate that the TrR signature is capable to guide repurposing drugs that were initially developed for other designations. However, due to some limitations of CMAP, such as limited drug perturbation data, drug coverage and dosage-dependent conditions (Musa, Ghoraie et al., 2018), candidates predicted from CMAP might not necessarily be the optimal choice to enter clinical settings. For example, AUY992 and Ganetespib, both of which were effective Hsp90 inhibitors but not present in CMAP database, were widely used to target the TrR population in clinical trials (6 trials in total up until 2019), whereas Geldanamycin, which was predicted and proven to effectively target this cohort via inhibiting the same pathway, was not found in clinical studies. We believe that valid drug prediction will be greatly improved with the evolution of future databases integrating both a CRISPR/Cas9 library for functional genomics and drug sensitivity gene expression profiles with a better spectrum of drug selections.

Although our TrR signature was able to predict drug candidates that target HDAC pathways or Hsp90 pathways, inhibitors targeting either pathway have proven difficult to utilize. HDAC inhibitors were usually found lacking tissue and drug specificity, which often lead to off-target effects (Clawson, 2016). Inhibitors of Hsp90, a chaperone heavily involved in post-translational modifications, frequently interferes with the majority of its targets, which could further disturb the functions of both tumor suppressors and oncogenes when targeting cancer cells (Soga, 2013). In our study, the network of genes within the TrR signature indicates possible mechanisms of action that TrR tumors inherit less cytotoxic T cell infiltration and suggests a new approach allowing for interrogation of immune modulation, such as a combination of IFN-γ and trastuzumab since the modulation of IFN-γ secretion was triggered by trastuzumab in the TrS cohort but not the TrR cohort. Despite substantial efforts made by other studies where immune-related signatures were associated with a response to trastuzumab-containing chemotherapy, their ability to predict benefit from specific HER2-targeted therapies and the *in vivo* effects of trastuzumab on immune response has not yet been elucidated. This limitation has led to another important aspect of our study - the utility of our archived in-house patient treatment naïve FFPE samples of patient core biopsies for initial diagnosis, which is the best timing for application of the TrR signature for clinical treatment decision.

Overall, our study suggests the importance of genomic evaluation beyond HER2 IHC/FISH testing from a systems biology approach, identified with gene expression. Our TrR signature accurately predicts response to trastuzumab *in vitro* and *in vivo*, and hopefully, can soon be used in clinical settings. Our study also suggests new approaches for therapeutic intervention to overcome trastuzumab resistance, such as increasing cytotoxic T cell infiltration by regulating several important cytokines or a regime of IFN-γ and trastuzumab combination in early stage disease, ideally in the preoperative setting. We believe that with our findings and appropriate settings in clinical trials, we will be able to define the optimal approach for each patient to make this disease finally curable.

## Materials and Methods

### Cell lines and selection

Human breast cancer cell lines BT474 (American Type Culture Collection (ATCC)) were exposed continuously to either 0.2 or 1μM Trastuzumab for 3 weeks when the survived formation of small clusters (BT474_R_0.2 and BT474_R_1, respectively, where BT474_R_1 was labeled as BT474_R sometimes) started to repropagate after the initial growth inhibition. The cell clusters were further cultured in the continuous presence of trastuzumab for an extra month. BT474 and the resistant clones were further maintained in DMEM (Corning) supplemented with 10% FBS (GE Healthcare-Hyclone). Additionally, the breast cancer cell lines T47D (ATCC), HCC1954 (ATCC) and ZR75-30 (ATCC) were cultured in RPMI (Gibco) media, supplemented with 10% FBS.

### Pipeline of developing signature and signature score

Publicly available transcriptome profile (GSE15043) of genetically engineered isogenic TrR lines of BT474 are used to generate the TrR signature using Bioconductor in R. Briefly, raw data were log2 transformed, quantile-normalized, and median polished. To determine a list of differentially expressed genes between BT474 and its derived resistant cell lines, we optimized the algorithm to maximize the accuracy of prediction while minimizing number of genes of interest in the desired samples by adjusting p-values (p = 0.01) and fold change (FC = 2) threshold. Performance of TrR signature was tested on independent in vitro and in vivo transcriptome datasets using Receiving Operation Curve (ROC). Signature scores were determined by calculating the correlations coefficient between medium centered gene expression levels within the signature and gene expression levels for that gene within a given sample, following quantile normalization.

### Patient survival analysis

Cohorts of HER2-positive breast cancer patients and HER2-positive gastric cancer patients were used as independent testing sets. Patient clinical data and RNAseq data were acquired from The Cancer Genome Atlas Program (TCGA). TrR signature scores were calculated following quantile normalization. To generate Kaplan-Meier curves, patients were divided based on the maximization of statistical difference in signature scores between the two groups.

### Cell line-derived xenografts

All animal procedures were conducted in compliance with National Institute of Health guidelines for animal research and approved by Institutional Animal Care and Use Committee (IACUC) at Aurora Research Institute. Five-week-old female Crl:Nu(NCr)-Foxn1nu nude mice were purchased from Charles River Laboratories and housed for another week in a specific pathogen-free environment, followed by subcutaneous injection into mammary fat pads with 1⨯10^6^ cells suspended in 100μl ice-cold Matrigel (Corning) at a 1:1 ratio for each mouse (n=10/each group). Tumor volumes, calculated by V = (length ⨯Width^2^)/2, and body weight were measured twice per week during treatment. Tumors dissected at the end point of each experiment were subjected to RNA extraction per protocol provided by manufacturer (Qiagen), followed by quantitative PCR assays for 43 genes in TrR signature.

### Quantitative PCR

RNAs of cells or tissues were extracted using RNeasy mini kit (Qiagen). Extracted RNA was further subjected to NanoDrop for RNA quantification. An iScript cDNA synthesis kit (BioRad) was used for cDNA synthesis from the extracted RNA. Quantitative PCR was prepared in biological triplicate using ITAQ(tm) Universal SYBR® Green Supermix (BioRad) and was performed by Roche Light Cycler 480 II instrument (Roche, NJ). Predesigned primers for genes (Integrated DNA Technologies, IDT) listed in TrR signature were optimized and details were listed in supplementary table 1. B2M (IDT, Assay ID: Hs.PT.58v.18759587) were used in the assay as an internal control. All gene expression levels were normalized to the level of B2M expression within the same sample, which is the internal standard and mRNA expression level calculated with the 2^−ΔΔCt^ method.

### Flow cytometry

For cell cycle analysis, cells were harvested and washed three times with PBS, followed by incubation in either PE-conjugated mouse anti-human IgG or PE-conjugated mouse anti-human HER2 antibody (1:500, BD) for one hour at room temperature. Samples were further subjected to flow cytometry and data were analyzed using Flowjo software.

### Dose response curves

Cell lines were plated at a density of 2000 cells per well (Nexcelom Bioscience Cellometer Auto 2000) in 96 well tissue culture plates using the appropriate culture medium, previously optimized from plate surface area and time of treatment incubation. Trastuzumab (SelleckChem) were serially diluted and the treated cells were incubated at 37 °C for 5 days, at which point the PrestoBlue cell viability assay (Invitrogen) was performed to determine relative cell population on a Synergy H1 microplate reader with Gen5 software (BioTek). The experiments were conducted in experimental triplicate, and the measurement was normalized to that of untreated cells.

### Prediction of agents targeting TrR tumors via Connectivity Map

In order to predict drugs that may effectively target TrR cells, we used Connectivity Map (CMAP), we enquired genes in the TrR signature with the fold change between BT474 TrR cell lines and BT474 parental lines (https://portals.broadinstitute.org/cmap/). For each gene in our TrR signature, we identified the corresponding feature set ID from approximately 20,000 probe sets on the HG-U133A array that are recognized by CMAP. These IDs were collated into tag set of “up” or “down”-regulated genes. Outcomes were reported as connectivity scores ranging from +1 to −1, where positive scores denote increased similarity while negative scores inverse similarity between the expression patterns induced by agents in CMAP and the gene profiles of reference samples.

### Quantification of soluble factors by ELISA

BT474 and BT474_R were seeded at 1×10^6^ cells in 6-well tissue culture plates and allowed to adhere overnight. Cells were then either treated with 1 μM PBS control or 1μM trastuzumab in 2 mL of growth media for 48 hours. The supernatants were collected, and the particulates removed by centrifugation. A Human XL Cytokine Array kit (R&D Systems) was used to measured cytokine secretion from tumor cells. Samples were prepared and the ELISAs were processed in accordance with the manufacturer’s instructions. After reference spot normalization, duplicate spots were averaged, followed by subtraction of an averaged background signal. Fold changes between corresponding signals of BT474_R versus BT474, both cell lines with versus without trastuzumab treatment were calculated and cross compared.

### Western blot

1⨯10^6^ cells were plated in 6-well cell culture dish (Corning), collected and lysed in RIPA buffer (Abcam) with protease inhibitors (Cell Signaling Technologies (CST)). Lysates were run in 4-12% Criterion Tris-HCL Protein gels (BioRad) in a Criterion Cell electrophoresis system (BioRad), followed by being transferred to Nitrocellulose, 0.2 μm membranes. Primary antibodies were applied at various dilutions: Actin (1: 1000, Sigma-Aldrich, St. Louis, MO), Beta-tubulin (1:500, CST), phospho-Histone H2A.X (Ser139) (1:1000, γH2AX; Millipore, Burlington, MA), phosho-Histone H3 (Ser10) (1: 500, CST), HER2/ErbB2 (1: 500, CST), MUC1(1:500, CST), Cleaved Caspase-3 (Asp175) (1: 1000, CST), and phospho-CDC2 (Y15) (1: 500, CST). The blots were then incubated with HRP-conjugated mouse (Santa Cruz Biotechnology (SCB)) or rabbit IgG antibodies at 1:1000 for one hour and the signals were detected on a LiCor3000 instrument (Li-Cor) using Western Clarity ECL (BioRad). Signals were measured and normalized, and experiments were repeated in triplicates.

### Clinical sample selection and Immunohistochemistry

From a single coordinated 15-hospital healthcare organization, medical records of patients diagnosed with HER2-positive breast cancer from 2007-2017 were assessed. Ten pairs of TrR tumors and TrS tumors were provided by our in-house Biorepository and Specimen Resource Center (BSRC), where the treatment naïve tumors from patients’ core biopsies were archived. Tumors of patients who responded to trastuzumab treatment and were relapse-free were classified as TrS tumors and tumors of patients who did not respond to trastuzumab regimen or who relapsed even though some clinical response were observed initially were classified as TrR tumors. ER status was matched between TrR tumors and TrS tumors when sample selections were made. Tissue sections were further dehydrated with Xylene and stained using EnVision Detection SystemsPeroxidase/DAB, Rabbit/Mouse kit (Agilent) kit per protocol previously described (Sun, Fang et al., 2017). Briefly, tissue sections with primary antibodies: CD4(1:20, Abcam), CD4 (1:10, Abcam) and MUC1(1: 500, CST), were incubated overnight at 4°C, followed by 1-hour incubation with Labelled Polymer-HRP at room temperature. Negative controls were treated identically, but without primary antibody. Subsequently, slides were incubated with DAB+ Chromogen, followed by counterstaining with hematoxylin. Pictures of slides after mounting with Permount (TFS) were captured under microscope (Olympus). Scores of IHC for all markers are calculated by the percentage of tumor cells multiplied by the intensity of markers.

### Statistical analysis

Data were analyzed by Student’s t-test and one- or two-way ANOVA per experiment design. A two-way ANOVA was used to assess the effects and interactions of two variables and multiple comparisons were achieved using Bonferroni’s *post hoc* test. All statistical analysis was completed using GraphPad Prism 6 and results are presented as mean ± SD of three independent experiments. Significance for normally distributed data was determined by student t-test (two groups) or by ANOVA with appropriate post-hoc tests. For data not normally distributed, a rank-sum test (two groups) or a Kruskal-Wallis test with an appropriate post-hoc test was used.

## Data Availability

The authors confirm that the data supporting the findings of this study are available within the article and its supplementary materials.

## Acknowledgements

This study is supported by Vince Lombardi Cancer Foundation and Aurora Health Care Seed Grant, Wisconsin, USA. We thank the Department of Biorepository at Aurora Research Institute for patient sample collection. The results shown in this study are partly based upon data generated by TCGA Research Network: http://cancergenome.nih.gov/. The networks represented in this study were generated through the use of QIAGEN’s Ingenuity Pathway Analysis (IPA®, QIAGEN Redwood City, www.qiagen.com/ingenuity). A part of the studies represented in this manuscript was obtained from RPPA database (funded by NCI #CA16672) and the Geodatabase (http://www.ncbi.nlm.nih.gov/geo/).

## Author Contributions

J. Yin designed, conceived, performed the experiments, analyzed the data and wrote the manuscript. A. Sand, A.T. Duffin, G.T Riddell, M. Piacsek, and C.Y. Sun performed the experiments, analyzed data and edited the manuscript. R.A. Rovin provided expertise and feedback. J.A. Tjoe provided clinical expertise and coordinated the project and provided feedback on the manuscript.

## References

Adams S, Gray RJ, Demaria S, Goldstein L, Perez EA, Shulman LN, Martino S, Wang M, Jones VE, Saphner TJ, Wolff AC, Wood WC, Davidson NE, Sledge GW, Sparano JA, Badve SS (2014) Prognostic value of tumor-infiltrating lymphocytes in triple-negative breast cancers from two phase III randomized adjuvant breast cancer trials: ECOG 2197 and ECOG 1199. J Clin Oncol 32: 2959–66

Arnaout AH, Dawson PM, Soomro S, Taylor P, Theodorou NA, Feldmann M, Fendly BM, Shepard HM, Shousha S (1992) HER2 (c-erbB-2) oncoprotein expression in colorectal adenocarcinoma: an immunohistological study using three different antibodies. J Clin Pathol 45: 726–7

Arteaga CL, Sliwkowski MX, Osborne CK, Perez EA, Puglisi F, Gianni L (2011) Treatment of HER2-positive breast cancer: current status and future perspectives. Nat Rev Clin Oncol 9: 16–32

Barretina J, Caponigro G, Stransky N, Venkatesan K, Margolin AA, Kim S, Wilson CJ, Lehar J, Kryukov GV, Sonkin D, Reddy A, Liu M, Murray L, Berger MF, Monahan JE, Morais P, Meltzer J, Korejwa A, Jane-Valbuena J, Mapa FA et al. (2012) The Cancer Cell Line Encyclopedia enables predictive modelling of anticancer drug sensitivity. Nature 483: 603–7

Basu A, Bodycombe NE, Cheah JH, Price EV, Liu K, Schaefer GI, Ebright RY, Stewart ML, Ito D, Wang S, Bracha AL, Liefeld T, Wawer M, Gilbert JC, Wilson AJ, Stransky N, Kryukov GV, Dancik V, Barretina J, Garraway LA et al. (2013) An interactive resource to identify cancer genetic and lineage dependencies targeted by small molecules. Cell 154: 1151–1161

Castagnoli L, Iezzi M, Ghedini GC, Ciravolo V, Marzano G, Lamolinara A, Zappasodi R, Gasparini P, Campiglio M, Amici A, Chiodoni C, Palladini A, Lollini PL, Triulzi T, Menard S, Nanni P, Tagliabue E, Pupa SM (2014) Activated d16HER2 homodimers and SRC kinase mediate optimal efficacy for trastuzumab. Cancer Res 74: 6248–59

Centis F, Tagliabue E, Uppugunduri S, Pellegrini R, Martignone S, Mastroianni A, Menard S, Colnaghi MI (1992) p185 HER2/neu epitope mapping with murine monoclonal antibodies. Hybridoma 11: 267–76

Clawson GA (2016) Histone deacetylase inhibitors as cancer therapeutics. Ann Transl Med 4: 287

Costello JC, Heiser LM, Georgii E, Gonen M, Menden MP, Wang NJ, Bansal M, Ammad-ud-din M, Hintsanen P, Khan SA, Mpindi JP, Kallioniemi O, Honkela A, Aittokallio T, Wennerberg K, Community ND, Collins JJ, Gallahan D, Singer D, Saez-Rodriguez J et al. (2014) A community effort to assess and improve drug sensitivity prediction algorithms. Nat Biotechnol 32: 1202–12

Damaskos C, Garmpis N, Valsami S, Kontos M, Spartalis E, Kalampokas T, Kalampokas E, Athanasiou A, Moris D, Daskalopoulou A, Davakis S, Tsourouflis G, Kontzoglou K, Perrea D, Nikiteas N, Dimitroulis D (2017) Histone Deacetylase Inhibitors: An Attractive Therapeutic Strategy Against Breast Cancer. Anticancer Res 37: 35–46

De Mattos-Arruda L, Cortes J (2012) Breast cancer and HSP90 inhibitors: is there a role beyond the HER2-positive subtype? Breast 21: 604–7

Digklia A, Wagner AD (2016) Advanced gastric cancer: Current treatment landscape and future perspectives. World J Gastroenterol 22: 2403–14

Dowsett M, Cooke T, Ellis I, Gullick WJ, Gusterson B, Mallon E, Walker R (2000) Assessment of HER2 status in breast cancer: why, when and how? Eur J Cancer 36: 170–6

Galluzzi L, Buque A, Kepp O, Zitvogel L, Kroemer G (2017) Immunogenic cell death in cancer and infectious disease. Nat Rev Immunol 17: 97–111

Gennari R, Menard S, Fagnoni F, Ponchio L, Scelsi M, Tagliabue E, Castiglioni F, Villani L, Magalotti C, Gibelli N, Oliviero B, Ballardini B, Da Prada G, Zambelli A, Costa A (2004) Pilot study of the mechanism of action of preoperative trastuzumab in patients with primary operable breast tumors overexpressing HER2. Clin Cancer Res 10: 5650–5

Gu L, Waliany S, Kane SE (2009) Darpp-32 and its truncated variant t-Darpp have antagonistic effects on breast cancer cell growth and herceptin resistance. PLoS One 4: e6220

Harris LN, You F, Schnitt SJ, Witkiewicz A, Lu X, Sgroi D, Ryan PD, Come SE, Burstein HJ, Lesnikoski BA, Kamma M, Friedman PN, Gelman R, Iglehart JD, Winer EP (2007) Predictors of resistance to preoperative trastuzumab and vinorelbine for HER2-positive early breast cancer. Clin Cancer Res 13: 1198–207

Herrmann F, Lehr HA, Drexler I, Sutter G, Hengstler J, Wollscheid U, Seliger B (2004) HER-2/neu-mediated regulation of components of the MHC class I antigen-processing pathway. Cancer Res 64: 215–20

Horm TM, Bitler BG, Broka DM, Louderbough JM, Schroeder JA (2012) MUC1 drives c-Met-dependent migration and scattering. Mol Cancer Res 10: 1544–54

Ibrahim EM, Al-Foheidi ME, Al-Mansour MM, Kazkaz GA (2014) The prognostic value of tumor-infiltrating lymphocytes in triple-negative breast cancer: a meta-analysis. Breast Cancer Res Treat 148: 467–76

Lamb J, Crawford ED, Peck D, Modell JW, Blat IC, Wrobel MJ, Lerner J, Brunet JP, Subramanian A, Ross KN, Reich M, Hieronymus H, Wei G, Armstrong SA, Haggarty SJ, Clemons PA, Wei R, Carr SA, Lander ES, Golub TR (2006) The Connectivity Map: using gene-expression signatures to connect small molecules, genes, and disease. Science 313: 1929–35

Loi S, Michiels S, Salgado R, Sirtaine N, Jose V, Fumagalli D, Kellokumpu-Lehtinen PL, Bono P, Kataja V, Desmedt C, Piccart MJ, Loibl S, Denkert C, Smyth MJ, Joensuu H, Sotiriou C (2014) Tumor infiltrating lymphocytes are prognostic in triple negative breast cancer and predictive for trastuzumab benefit in early breast cancer: results from the FinHER trial. Ann Oncol 25: 1544–50

Loi S, Sirtaine N, Piette F, Salgado R, Viale G, Van Eenoo F, Rouas G, Francis P, Crown JP, Hitre E, de Azambuja E, Quinaux E, Di Leo A, Michiels S, Piccart MJ, Sotiriou C (2013) Prognostic and predictive value of tumor-infiltrating lymphocytes in a phase III randomized adjuvant breast cancer trial in node-positive breast cancer comparing the addition of docetaxel to doxorubicin with doxorubicin-based chemotherapy: BIG 02-98. J Clin Oncol 31: 860–7

Loibl S, de la Pena L, Nekljudova V, Zardavas D, Michiels S, Denkert C, Rezai M, Bermejo B, Untch M, Lee SC, Turri S, Urban P, Kummel S, Steger G, Gombos A, Lux M, Piccart MJ, Von Minckwitz G, Baselga J, Loi S (2017) Neoadjuvant buparlisib plus trastuzumab and paclitaxel for women with HER2+ primary breast cancer: A randomised, double-blind, placebo-controlled phase II trial (NeoPHOEBE). Eur J Cancer 85: 133–145

Luen SJ, Salgado R, Fox S, Savas P, Eng-Wong J, Clark E, Kiermaier A, Swain SM, Baselga J, Michiels S, Loi S (2017) Tumour-infiltrating lymphocytes in advanced HER2-positive breast cancer treated with pertuzumab or placebo in addition to trastuzumab and docetaxel: a retrospective analysis of the CLEOPATRA study. Lancet Oncol 18: 52–62

Maimela NR, Liu S, Zhang Y (2019) Fates of CD8+ T cells in Tumor Microenvironment. Comput Struct Biotechnol J 17: 1–13

Mani A, Roda J, Young D, Caligiuri MA, Fleming GF, Kaufman P, Brufsky A, Ottman S, Carson WE, 3rd, Shapiro CL (2009) A phase II trial of trastuzumab in combination with low-dose interleukin-2 (IL-2) in patients (PTS) with metastatic breast cancer (MBC) who have previously failed trastuzumab. Breast Cancer Res Treat 117: 83–9

McClory S, Hughes T, Freud AG, Briercheck EL, Martin C, Trimboli AJ, Yu J, Zhang X, Leone G, Nuovo G, Caligiuri MA (2012) Evidence for a stepwise program of extrathymic T cell development within the human tonsil. J Clin Invest 122: 1403–15

Muller P, Kreuzaler M, Khan T, Thommen DS, Martin K, Glatz K, Savic S, Harbeck N, Nitz U, Gluz O, von Bergwelt-Baildon M, Kreipe H, Reddy S, Christgen M, Zippelius A (2015) Trastuzumab emtansine (T-DM1) renders HER2+ breast cancer highly susceptible to CTLA-4/PD-1 blockade. Sci Transl Med 7: 315ra188

Musa A, Ghoraie LS, Zhang SD, Glazko G, Yli-Harja O, Dehmer M, Haibe-Kains B, Emmert-Streib F (2018) A review of connectivity map and computational approaches in pharmacogenomics. Brief Bioinform 19: 506–523

Musolino A, Naldi N, Bortesi B, Pezzuolo D, Capelletti M, Missale G, Laccabue D, Zerbini A, Camisa R, Bisagni G, Neri TM, Ardizzoni A (2008) Immunoglobulin G fragment C receptor polymorphisms and clinical efficacy of trastuzumab-based therapy in patients with HER-2/neu-positive metastatic breast cancer. J Clin Oncol 26: 1789–96

Okita R, Mougiakakos D, Ando T, Mao Y, Sarhan D, Wennerberg E, Seliger B, Lundqvist A, Mimura K, Kiessling R (2012) HER2/HER3 signaling regulates NK cell-mediated cytotoxicity via MHC class I chain-related molecule A and B expression in human breast cancer cell lines. J Immunol 188: 2136–45

Perez EA, Ballman KV, Tenner KS, Thompson EA, Badve SS, Bailey H, Baehner FL (2016) Association of Stromal Tumor-Infiltrating Lymphocytes With Recurrence-Free Survival in the N9831 Adjuvant Trial in Patients With Early-Stage HER2-Positive Breast Cancer. JAMA Oncol 2: 56–64

Perez EA, Romond EH, Suman VJ, Jeong JH, Davidson NE, Geyer CE, Jr., Martino S, Mamounas EP, Kaufman PA, Wolmark N (2011) Four-year follow-up of trastuzumab plus adjuvant chemotherapy for operable human epidermal growth factor receptor 2-positive breast cancer: joint analysis of data from NCCTG N9831 and NSABP B-31. J Clin Oncol 29: 3366–73

Perez EA, Romond EH, Suman VJ, Jeong JH, Sledge G, Geyer CE, Jr., Martino S, Rastogi P, Gralow J, Swain SM, Winer EP, Colon-Otero G, Davidson NE, Mamounas E, Zujewski JA, Wolmark N (2014) Trastuzumab plus adjuvant chemotherapy for human epidermal growth factor receptor 2-positive breast cancer: planned joint analysis of overall survival from NSABP B-31 and NCCTG N9831. J Clin Oncol 32: 3744–52

Pochampalli MR, Bitler BG, Schroeder JA (2007) Transforming growth factor alpha dependent cancer progression is modulated by Muc1. Cancer Res 67: 6591–8

Repka T, Chiorean EG, Gay J, Herwig KE, Kohl VK, Yee D, Miller JS (2003) Trastuzumab and interleukin-2 in HER2-positive metastatic breast cancer: a pilot study. Clin Cancer Res 9: 2440–6

Salgado R, Denkert C, Campbell C, Savas P, Nuciforo P, Aura C, de Azambuja E, Eidtmann H, Ellis CE, Baselga J, Piccart-Gebhart MJ, Michiels S, Bradbury I, Sotiriou C, Loi S (2015) Tumor-Infiltrating Lymphocytes and Associations With Pathological Complete Response and Event-Free Survival in HER2-Positive Early-Stage Breast Cancer Treated With Lapatinib and Trastuzumab: A Secondary Analysis of the NeoALTTO Trial. JAMA Oncol 1: 448–54

Singh JC, Jhaveri K, Esteva FJ (2014) HER2-positive advanced breast cancer: optimizing patient outcomes and opportunities for drug development. Br J Cancer 111: 1888–98

Slamon DJ, Clark GM, Wong SG, Levin WJ, Ullrich A, McGuire WL (1987) Human breast cancer: correlation of relapse and survival with amplification of the HER-2/neu oncogene. Science 235: 177–82

Soga S (2013) [Drug discovery and research of heat shock protein 90 (Hsp90) inhibitor]. Nihon Yakurigaku Zasshi 141: 9–14

Stagg J, Loi S, Divisekera U, Ngiow SF, Duret H, Yagita H, Teng MW, Smyth MJ (2011) Anti-ErbB-2 mAb therapy requires type I and II interferons and synergizes with anti-PD-1 or anti-CD137 mAb therapy. Proc Natl Acad Sci U S A 108: 7142–7

Stanton SE, Disis ML (2016) Clinical significance of tumor-infiltrating lymphocytes in breast cancer. J Immunother Cancer 4: 59

Sun C, Fang Y, Yin J, Chen J, Ju Z, Zhang D, Chen X, Vellano CP, Jeong KJ, Ng PK, Eterovic AKB, Bhola NH, Lu Y, Westin SN, Grandis JR, Lin SY, Scott KL, Peng G, Brugge J, Mills GB (2017) Rational combination therapy with PARP and MEK inhibitors capitalizes on therapeutic liabilities in RAS mutant cancers. Sci Transl Med 9

Triulzi T, De Cecco L, Sandri M, Prat A, Giussani M, Paolini B, Carcangiu ML, Canevari S, Bottini A, Balsari A, Menard S, Generali D, Campiglio M, Di Cosimo S, Tagliabue E (2015) Whole-transcriptome analysis links trastuzumab sensitivity of breast tumors to both HER2 dependence and immune cell infiltration. Oncotarget 6: 28173–82

Varadan V, Gilmore H, Miskimen KL, Tuck D, Parsai S, Awadallah A, Krop IE, Winer EP, Bossuyt V, Somlo G, Abu-Khalaf MM, Fenton MA, Sikov W, Harris LN (2016) Immune Signatures Following Single Dose Trastuzumab Predict Pathologic Response to PreoperativeTrastuzumab and Chemotherapy in HER2-Positive Early Breast Cancer. Clin Cancer Res 22: 3249–59

Varchetta S, Gibelli N, Oliviero B, Nardini E, Gennari R, Gatti G, Silva LS, Villani L, Tagliabue E, Menard S, Costa A, Fagnoni FF (2007) Elements related to heterogeneity of antibody-dependent cell cytotoxicity in patients under trastuzumab therapy for primary operable breast cancer overexpressing Her2. Cancer Res 67: 11991–9

Wongtrakoongate P (2015) Epigenetic therapy of cancer stem and progenitor cells by targeting DNA methylation machineries. World J Stem Cells 7: 137–48

Zhang N, Bevan MJ (2011) CD8(+) T cells: foot soldiers of the immune system. Immunity 35: 161–8

Zhong H, Davis A, Ouzounova M, Carrasco RA, Chen C, Breen S, Chang YS, Huang J, Liu Z, Yao Y, Hurt E, Moisan J, Fung M, Tice DA, Clouthier SG, Xiao Z, Wicha MS, Korkaya H, Hollingsworth RE (2016) A Novel IL6 Antibody Sensitizes Multiple Tumor Types to Chemotherapy Including Trastuzumab-Resistant Tumors. Cancer Res 76: 480–90

